# Health Economics Assessment of Statin Therapy Initiation Thresholds for Atherosclerosis Prevention in China: a Cost-Effectiveness Analysis

**DOI:** 10.1101/2023.08.03.23293584

**Authors:** Tianyu Feng, Xiaolin Zhang, Jiaying Xu, Shang Gao, Xihe Yu

## Abstract

**Background:** The latest revised Chinese guidelines for the management of dyslipidemia have lowered the 10-year risk threshold for initiating statins for primary prevention of atherosclerotic heart disease. The aim of this study was to examine the negative impact of different statin treatment initiation thresholds on diabetes in a Chinese population and to assess their health economics value.

**Methods:** In this study, we constructed an event probability-based Microsimulation model to assess the health economics value of statin therapy. The model was based on the Prediction for atherosclerotic cardio-vascular disease (ASCVD) risk in China (China-PAR) and used data from a nationally representative survey and published meta-analysis of the Chinese middle-aged and elderly population as input. We evaluated four different strategies: a 7.5% 10-year risk threshold strategy, a guideline strategy, a 15% 10-year risk threshold strategy, and a 20% 10-year risk threshold strategy. Additionally, we calculated the incremental cost per quality-adjusted life year (QALY) obtained for each strategy to better understand the economics of the various strategies.

**Result:** The incremental cost per QALY for the 10% 10-year risk threshold strategy, compared to the untreated, was $52,218.75. The incremental cost per QALY for the guideline strategy, compared to the 7.5% 10-year risk threshold strategy, was $464,614.36. These results were robust in most sensitivity analyses.

**Conclusion:** The current 10-year ASCVD risk thresholds used in China’s dyslipidemia management guidelines are cost-effective in preventing ASCVD events and should be maintained with the current statin initiation thresholds. Relaxing the initiation threshold as willingness to pay increases would be more cost-effective.

## Introduction

The disease spectrum in China has undergone significant changes in the last three decades. Cardiovascular diseases have replaced infectious diseases as the leading cause of death[1]. According to statistics, more than 40% of deaths in China are due to cardiovascular diseases, with dyslipidemia being an important risk factor for atherosclerotic cardiovascular disease (ASCVD)[2, 3]. To reduce the risk of major cardiovascular adverse events, clinical guidelines from various countries have recommended the use of statins to manage dyslipidemia[4–6]. A mixture of risk factor rules and absolute risk rules are used in the vast majority of guidelines to determine the conditions that trigger statin intervention, but the threshold for statin initiation recommended in different guidelines varies widely. For example, in the 2016 guidelines in China, statins are recommended for primary prevention when a patient has a 10-year ASCVD risk of 10%[4], whereas in Scotland, intervention is triggered when a patient has a 10-year ASCVD risk of 20%[7]. Some countries have set this standard even lower, such as the American Heart Association in its 2018 guideline on blood cholesterol management and the National Institute for Health and Clinical Excellence (NICE) in the UK in the latest version of its revised guideline in 2023, which sets a 10-year incidence of ASCVD of 7.5% as the threshold for triggering statin intervention[5, 6]. An Australian study concluded that a 10-year risk threshold of 15% is more cost-effective than a risk threshold of 5% or 10% for patients with coronary heart disease (CHD) and stroke[8]. It is important to consider the economic value when developing intervention initiation thresholds, especially in developing countries such as China.

Chinese guidelines for dyslipidemia management recommend initiating statin therapy when a patient’s 10-year risk of ASCVD exceeds 10%. However, this threshold remains a subject of debate within the academic community[4, 9]. Economic considerations must be taken into account when setting the threshold. A threshold that is too low may result in increased treatment costs for less vulnerable subgroups without a corresponding increase in health benefits. Conversely, a threshold that is too high may result in undertreatment of patients who could benefit from statins. Although statins effectively control dyslipidemia, they also increase the user’s risk of developing diabetes, which significantly affects the occurrence of ASCVD[10–12]. With the highest burden of diabetes in the world[13], China needs to investigate the effect of different statin initiation thresholds on diabetes risk and analyze the cost-effectiveness of statins in preventing ASCVD. This study aims to provide valuable reference information for clinicians and policymakers to develop rational, effective, safe, and economically feasible dyslipidemia management programs. Each country must conduct a cost-effectiveness analysis to determine the optimal threshold for initiating treatment based on its population profile, healthcare level, and economic level.

## Methods

### Model Structure

This study developed a microsimulation model to simulate lifetime ASCVD-related outcomes and costs for individuals aged 40-89 years[14]. The model is based on a modified Markov state model, where individuals transition between health states based on transfer probabilities. Upon entering the model, each individual is assigned parameters such as age, sex, physiological indicators, disease history, and treatment status based on demographic distribution parameters obtained from the 2015 China Health and Retirement Longitudinal Study (CHARLS). The upper age limit of individuals in the model is 89 years old due to the small proportion of the Chinese population aged 90 and above[15]. Individual physiological parameters are updated according to a regression equation as a function of age, as described in Table S5 of the Supplementary Material.

The probabilities of lethal and nonlethal cardiovascular and stroke events for individuals in the model were calculated from the risk function constructed by China-PAR (Supplementary material Table S6) [9, 16]. The risk of ASCVD is recalculated for individuals in each cycle based on their current physiological parameters, medical history, and treatment status. In the model, individuals are also affected by background mortality. The probability of a diabetic event is based on the annual incidence of diabetes in Chinese adults obtained from previous studies, with the relative risk of statin-induced diabetes calculated and reassigned if the individual is taking statins. Individuals who discontinue treatment will have their baseline incidence rate restored in the next cycle.

The model runs on a one-year cycle, with cost and health utility values updated at the end of each cycle. If an individual does not die or reach the modeled age of simulation termination, the simulation proceeds to the next cycle. If an individual dies or reaches the age of simulation termination, the simulation stops and costs, health benefits, and occurrences of events are recorded throughout the simulation period. The model structure is shown in Figure 1 and was implemented using Treeage Pro2019 software.

**Figure 1.**
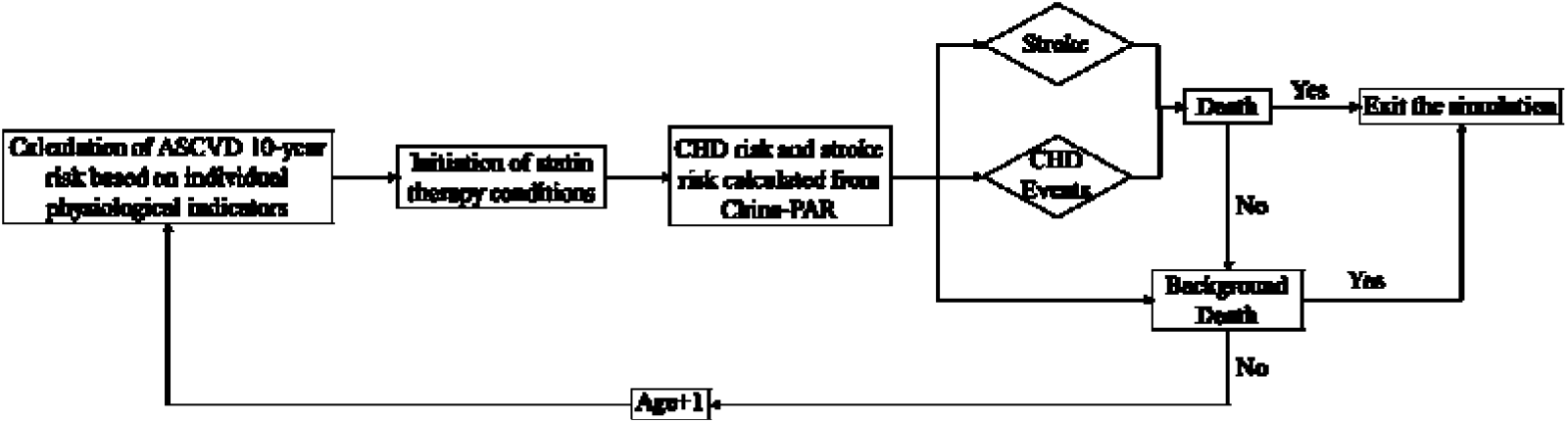
Structure of the microsimulation model. CHD events include all fatal coronary events and non-fatal AMI.

### Risk of ASCVD events

This study used a total risk equation derived from China-PAR to assess the 10-year composite risk of ASCVD events[9, 16]. The 2016 revised Chinese guidelines for the prevention and treatment of dyslipidemia in adults and the Chinese guidelines for cardiovascular disease prevention both recommend using China-PAR as an ASCVD risk assessment tool[4]. The China-PAR risk assessment tool has been widely used in clinical care in China and has been shown to be reliable and valid. China-PAR predicts fatal CHD events (all events caused by acute myocardial infarction (AMI) or other fatal events caused by coronary death), non-fatal AMI, and the 10-year composite probability of fatal and non-fatal stroke. The 10-year probability was converted to a 1-year probability using the DEALE method[17]. The 10-year composite probability of fatal AMI included the 10-year probability of CHD (29.46%) and stroke (70.54%), using their respective proportions of total CHD and stroke events[18]. Patients who survived a CHD event or stroke and did not have a recurrent CHD or stroke event had more than twice the background mortality rate at all follow-up cycles[19]. In the model, individuals who never experienced any ASCVD event were likely to die from background mortality (Supplementary material Tables S2).

### Simulation population

At the beginning of the simulation, the model consisted of 10,000 individuals without a history of ASCVD. Parameters for ASCVD-related risk factors were obtained from the 2015 CHARLS, and all other model inputs are shown in the Supplementary Material (Supplementary material Tables S1, S3 and S4)[20]. To model changes in ASCVD risk factors (mainly HDL-C, TC, SBP, and WC) over time, cross-sectional linear regression was performed using CHARLS data to obtain the relationship between ASCVD risk factors and age (Supplementary material Tables S5) in the China-PAR risk function[17]. Data on the prevalence of diabetes and the increased risk of developing diabetes due to statins at each age were obtained from studies in the literature[21].

### Treatment Strategies

The guideline treatment-initiation strategy is illustrated in electronic supplementary material Fig. S1. This study discusses statin treatment regimens based on the 2023 edition of the Chinese guidelines for lipid management, focusing on the 10-year ASCVD composite risk threshold in the guidelines. The China-PAR risk equation was used to determine patients’ 10-year risk of combined ASCVD onset[22]. The guidelines recommend the use of moderate-intensity statins as primary prevention. High-intensity statin therapy was not analyzed because the guidelines suggest that it should be used with caution in the Chinese population[4]. Based on previous studies, it was assumed that 40% of patients would discontinue treatment on their own after one year, and that all patients should adhere to long-term statin treatment[23, 24].

Since diabetes significantly affects the risk of developing combined ASCVD and statins can slightly increase the risk of developing type 2 diabetes[17, 25], this study focuses on the effect of statin-induced diabetes on the combined risk of ASCVD[26, 27]. It was assumed that patients with statin-induced diabetes had good glycemic control and no other comorbidities. In the simulation model, only the effects of statin-induced diabetes on quality of life and costs were included.

We analyzed 10-year risks of 7.5%, 10%, and 15% to explore optimal treatment initiation thresholds. These simulated regimens were similar to the strategies in the guidelines, with treatment initiation based on the China-PAR risk function. Treatment-free simulations were also performed.

### Cost and health utility

This study uses the perspective of the Chinese health care system, and costs include only direct medical costs associated with ASCVD, such as statin purchase, inpatient, and outpatient costs (Table 1). Drug costs were obtained based on the 2020 national negotiated prices. We followed the methodology of a similar study and calculated annual drug costs by averaging the lowest winning prices of statins from local government procurement catalogs in each region of China in the first quarter of 2020. Assuming that post-CHD and post-stroke individuals require one outpatient visit per year for the remainder of their lives, we expanded the costs to 2022 using the China Healthcare Component Consumer Price Index.

**Table 1.** Parameters used in the model.

| Parameter | Value | Distribution | Standard error | References |
| --- | --- | --- | --- | --- |
| Acute disutility |  |  |  | [28] |
| Coronary heart disease | 0.439 | Beta | 0.018 |  |
| Stroke | 0.92 | Beta | 0.04 |  |
| Post-event long-term disutility |  |  |  | [28,29] |
| Acute myocardial infarction | 0.107 | Beta | 0.019 |  |
| Stroke | 0.266 | Beta | 0.02 |  |
| Treatment effect on CHD (OR) | 0.7 | Log-normal | 0.072 | [22] |
| Treatment effect on stroke (OR) | 0.81 | Log-normal | 0.069 | [22] |
| Risk of statin-induced diabetes (OR) | 1.21 | Beta |  | [30] |
| Annual statin treatment costs | 1,149.75 | Gamma |  |  |
| Costs (2022 CN¥) | AMI | Stroke |  |  |
| Hospitalization costs | 22,611 | 13,983 | Gamma | [31] |
| First-year long-term costs | 5,255.85 | 2652 | Gamma | [32] |
| Office visit costs in subsequent years | 585.87 | 565 | Gamma | [33] |
| CHD mortality by age group | Men | Women |  | [32] |
| 34-44 | 0.12 | 0.18 | Beta |  |
| 45-54 | 0.21 | 0.23 | Beta |  |
| 55-64 | 0.29 | 0.27 | Beta |  |
| 65-74 | 0.33 | 0.43 | Beta |  |
| 75-84 | 0.48 | 0.51 | Beta |  |
| Stroke mortality by age group | Men | Women |  | [32] |
| 34-44 | 0.25 | 0.18 | Beta |  |
| 45-54 | 0.18 | 0.14 | Beta |  |
| 55-64 | 0.12 | 0.15 | Beta |  |
| 65-74 | 0.2 | 0.2 | Beta |  |
| 75-84 | 0.45 | 0.45 | Beta |  |

QALYs were calculated using age-specific utility weights and event-related negative utility weights, which can be further categorized into acute and long-term negative utility for patients surviving only in the acute phase(Table 1). Age-specific utility weights were derived from the EQ-5D survey of the Chinese general population (Supplementary material Tables S2)[34]. Acute and long-term negative utility weights were extracted from the Global Burden of Disease project and the Chinese EQ-5D survey of patients with chronic diseases[29]. In the baseline case, an annual discount rate of 3% was used[35].

### Intervention Strategy Comparisons

This study evaluates strategies using the incremental cost-benefit ratio (ICER). The cost-benefit ratio of each alternative is calculated, and inferior strategies are removed. China’s gross domestic product (GDP) per capita in 2022 (CNY85,700) is used as the cost-effectiveness criterion, and three times the GDP (CNY257,100) is used as the minimum criterion to be cost-effective[36]. In addition, this study employs cost-acceptability curves to illustrate the probability that each strategy is cost-effective at different levels of willingness-to-pay (WTP).

### Sensitivity Analysis

We performed one-way and probabilistic sensitivity analyses (PSA) on the model parameters to assess the robustness of the results when the parameters were varied. In the univariate sensitivity analysis, we focused on the effects of statin effectiveness, price, and induction of diabetes on the optimal ASCVD treatment threshold. In PSA, we assessed overall model uncertainty through 1000 randomized simulations.

### Model Validation

The model used in this study was adapted from an extensively validated Microsimulation model[14, 37], and its internal and external validity were validated based on the model output[38, 39]. Specifically, the internal validity of the model was validated by regression analysis, calculating the expected probabilities of CHD and stroke events for each sex-different age group in the untreated group model against the mean of the simulation results (Supplementary material Figure S2). External validity was verified by comparing simulated ASCVD incidence in the age-sex group (10-year interval) with results from other studies (Supplementary material Table S6)[40].

## Results

### Model calibration and validation

Fig S2 in supplementary material displays the internal validation results of the model. The coefficient of determination R2 for the linear regression on the mean of the 1-year expected probability of CHD and stroke events and the corresponding simulated values for the untreated group is 0.9278, indicating good internal validity. The simulated stroke incidence in Table 2 aligns well with the prospective study results. China’s current published life expectancy per capita is 75.4 years, while the untreated model simulates a life expectancy of 74.9 years, demonstrating good external validity.

**Table 2.**
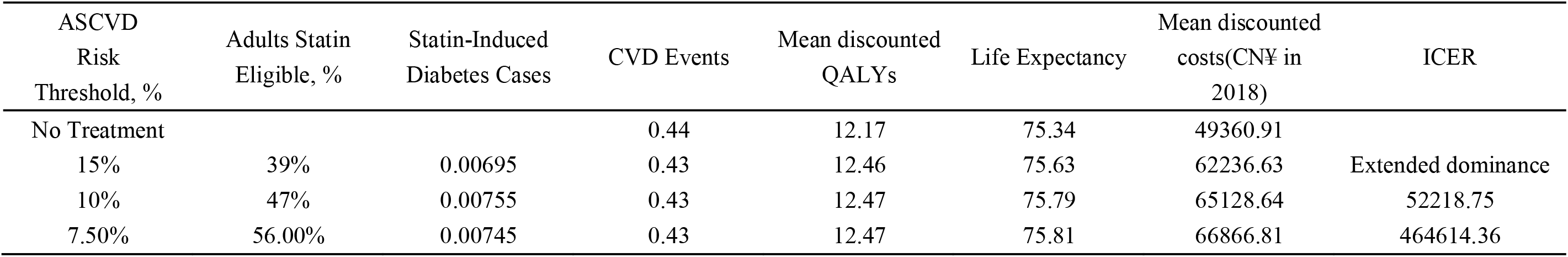
Base-case results of ICERs.

| ASCVD<br>Risk<br>Threshold, % | Adults Statin<br>Eligible, % | Statin-Induced<br>Diabetes Cases | CVD Events | Mean discounted<br>QALYs | Life Expectancy | Mean discounted<br>costs(CN¥ in<br>2018) | ICER |
| --- | --- | --- | --- | --- | --- | --- | --- |
| No Treatment |  |  | 0.44 | 12.17 | 75.34 | 49360.91 |  |
| 15% | 39% | 0.00695 | 0.43 | 12.46 | 75.63 | 62236.63 | Extended dominance |
| 10% | 47% | 0.00755 | 0.43 | 12.47 | 75.79 | 65128.64 | 52218.75 |
| 7.50% | 56.00% | 0.00745 | 0.43 | 12.47 | 75.81 | 66866.81 | 464614.36 |

### Clinical Event Results

An ASCVD threshold of 7.5% or higher was associated with an estimated 48% of adults being eligible for statin therapy. Strategies with higher ASCVD thresholds were linked to fewer cases of statin-induced diabetes but more CVD events. When applied to the 573 million adults aged 40 to 75 years in China, shifting from a 15% to a 10% ASCVD threshold was associated with an estimated 1,088,716 additional CVD events avoided. Shifting from a 10% to a 7.5% ASCVD threshold was associated with an estimated 687,610 additional CVD events avoided. In contrast, the number of statin-induced diabetes cases was small. This result demonstrates the net effect of statins in reducing cardiovascular events.

### Cost-effectiveness analysis

Table 2 presents the base case results of this study. The ICERs for the 15% risk threshold strategy compared to no treatment were $43,951.82/QALY. The 15% risk threshold strategy is the expansion advantage strategy and was removed from the analysis. The ICERs for the guideline strategy compared to the untreated were $52,218.75/QALY. The ICERs for the 7.5% risk threshold strategy compared to the guideline strategy were $464,614.36/QALY. Therefore, when using the 3-GDP criterion, the 7.5% risk threshold strategy was optimal among the three strategies. When using the 1-GDP criterion, the 10% risk threshold strategy was optimal among the three strategies.

### Sensitivity Analysis

The acceptability curves for the PSA results are displayed in the Figure 2. When using the 3-GDP cost-effectiveness criterion, the guidance strategy has a 32% probability of being optimal, while the 7.5% risk threshold strategy has a 62% probability. When using the 1-GDP criterion, the guidance strategy and the 7.5% risk threshold strategy have equal cost-effectiveness, both with a 30% probability of being optimal. The cost-acceptability curve suggests that the current guideline strategy is more cost-effective than the 7.5% risk threshold strategy when the WTP is below 1-GDP.

**Figure 2.**
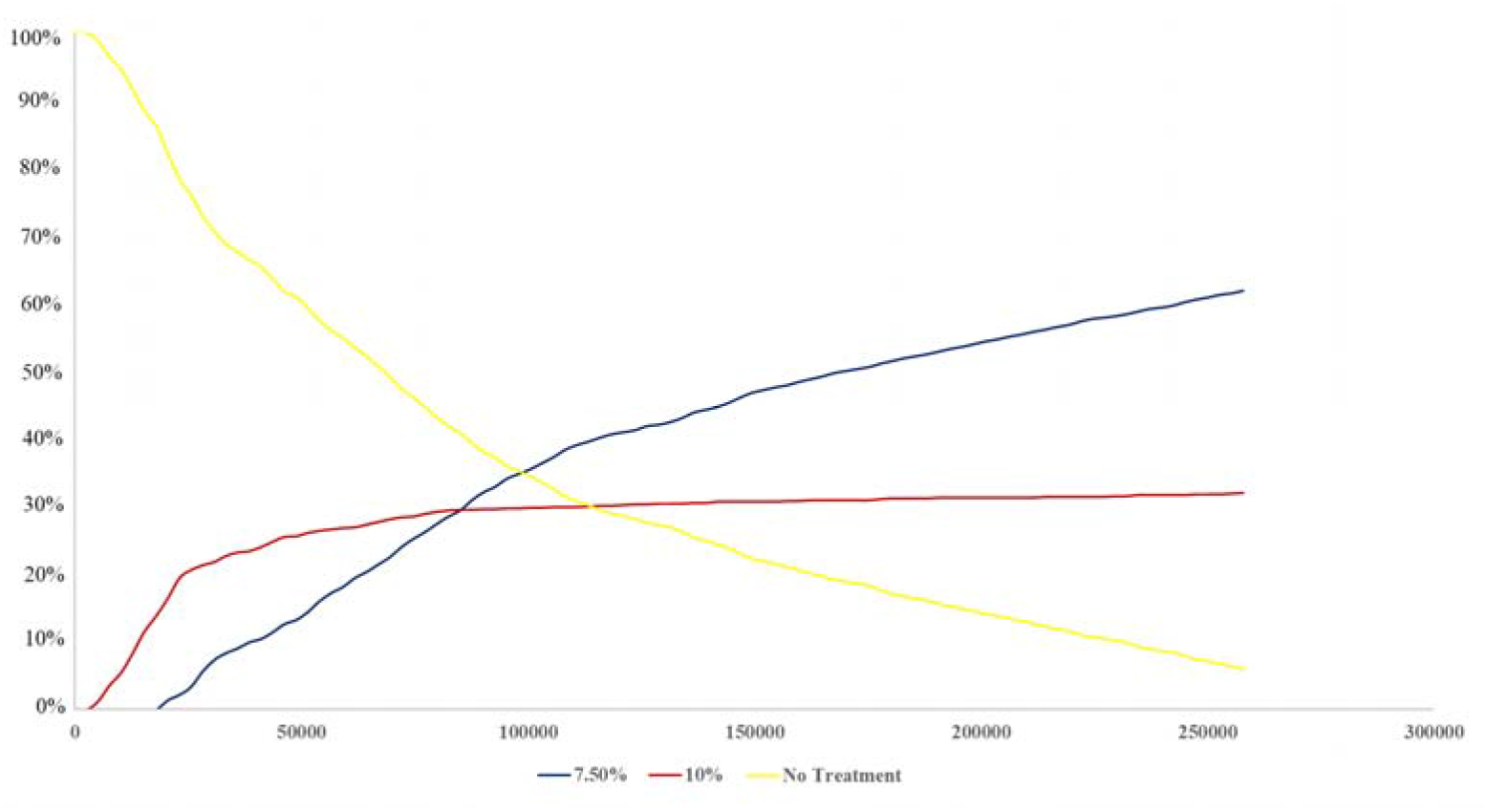
Acceptability curves were generated using probabilistic sensitivity analysis results. The horizontal coordinate of the figure represents willingness to pay, calculated in RMB.

The tornado plot displays the one-way sensitivity analysis results (figure 3). When all parameters are varied within their range of variability, the degree of variation in ICER remains within an acceptable range around 1-GDP. Changes in statin prices and the risk of statin-induced diabetes have the greatest impact on cost-effectiveness results.

**Figure 3.**
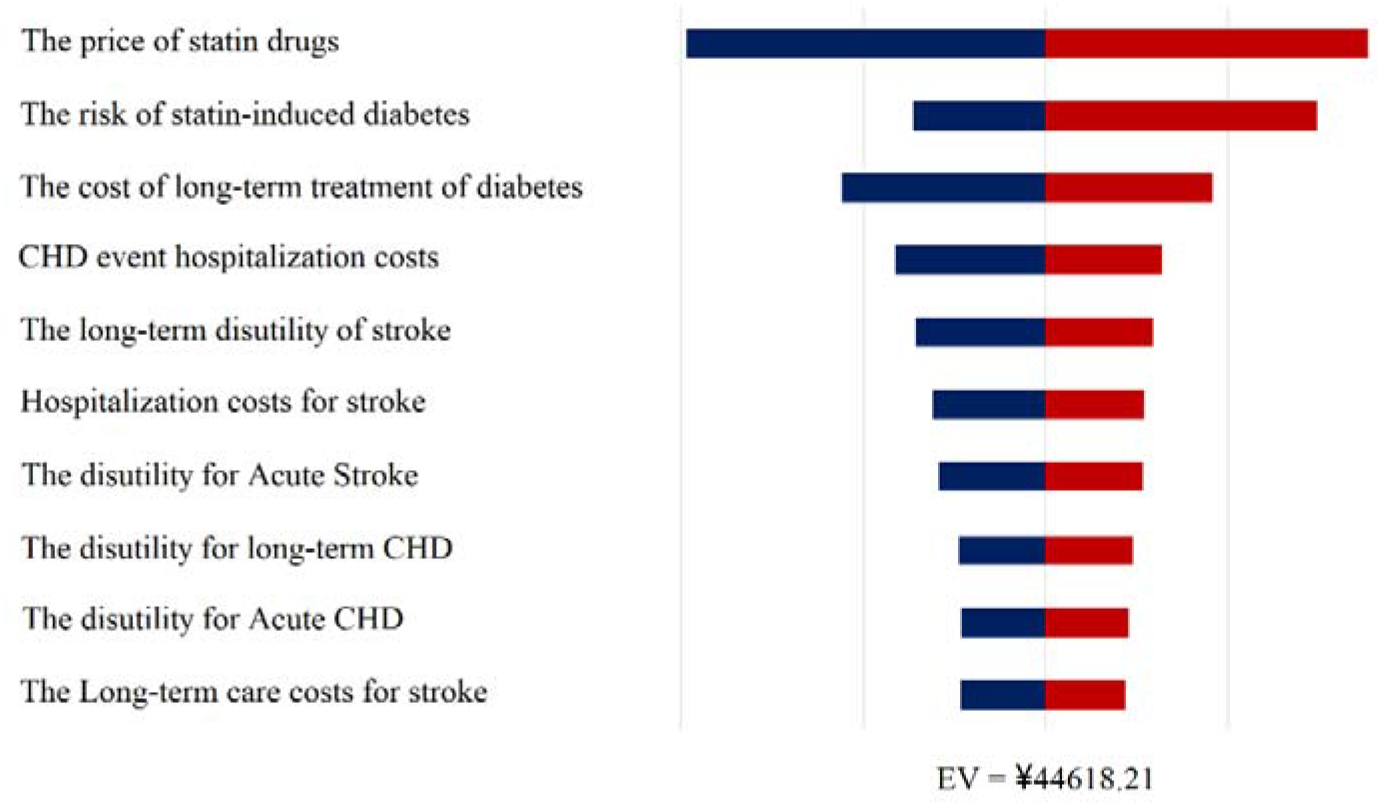
The tornado plot displays the results of the deterministic sensitivity analysis. The price of statins and the risk of statin-induced diabetes had the greatest impact on the results.

## Discussion

The primary goal of healthcare systems is to optimize health benefits for the population while minimizing expenditure, particularly in the health economics of preventative therapeutic measures[41]. Statins have been demonstrated to be effective in preventing ASCVD, yet their use has been associated with an increased incidence of diabetes, presenting a challenge for policymakers, healthcare professionals, and patients[42, 43]. Our analysis revealed that using a 7.5% 10-year ASCVD risk as the initiation threshold for statin prevention of ASCVD events was more effective than the current Chinese dyslipidemia management guidelines’ recommendation of a 10% 10-year ASCVD risk when 3-GDP was used as the criterion. Sensitivity analysis results indicated that the effects of statin sales price and the risk prevention strategy for statin-induced diabetes were more pronounced. Notably, when the price of statins is reduced by 20%, the ICER for the 7.5% threshold strategy will be less than 2-GDP. Overall, our findings suggest that the optimal treatment threshold varies depending on the cost-effectiveness threshold used.

For adults without cardiovascular disease, the decision to initiate statin therapy should be based on evidence-based evidence and patient preference. When a patient’s ASCVD risk reaches the initiation threshold, they may have valid personal reasons for declining statin treatment[44]. In China, financial reasons often lead patients to discontinue treatment. For many lower-income patients without well-established health insurance, the cost is prohibitive[45]. Our one-way sensitivity analysis indicated that statin-induced diabetes has a significant impact on simulation outcomes. Therefore, greater attention should be paid to the glycemic profile of patients taking statins and to their long-term normative glucose monitoring.

Studies evaluating the economics of statin clinical guidelines in other countries and regions have concluded that the threshold for initiating statin therapy could be further reduced. Pandya et al. assessed the cost-effectiveness of the 2013 ACC/AHA guidelines and alternative risk thresholds for preventing ASCVD events. They determined that the 10-year ASCVD ≥7.5% risk threshold used in the ACC/AHA guidelines was acceptably cost-effective, but that cost-effectiveness would be more pronounced if the 10-year ASCVD risk threshold were further reduced (e.g., to ≥3% or ≥4%)[14]. Linda J et al. analyzed current cardiovascular prevention guidelines in Australia and determined that the greatest cost-effectiveness was achieved by recommending statins for individuals with a five-year ASCVD risk greater than 5% or a 10-year ASCVD risk greater than 10%[8]. These studies suggest that more aggressive statin therapy initiation threshold strategies could be further explored. However, because these studies were primarily based on the demographics, healthcare levels, and economic development of developed countries, their results may not be replicable in developing countries like China. Specific statin initiation thresholds should consider the country’s willingness to pay.

Past economic studies of statins have not addressed the impact of diabetes, preventing the analysis from fully reflecting the actual outcomes of statins in preventing ASCVD. However, the risk functions used in past studies are not the parameters currently needed on the front line and are more difficult to obtain, causing the analysis results to deviate somewhat from the actual situation and making it challenging to meet the practical needs of grassroots work. As a result, previous studies have struggled to produce results that provide practical guidance. By improving the model structure and correcting the parameters, this study obtained valuable references for practical clinical work. Cobiac et al. used the average cost-effectiveness ratio of each strategy, rather than ICER, to compare different absolute risk threshold strategies[8]. Jiang et al. analyzed the cost-effectiveness of different risk thresholds using a DEM model based on a national survey[46]. Unlike our study, their study did not include the risk of statin-induced diabetes in the model, omitting an important factor for ASCVD prevention. Our study addressed this omission.

Based on this study’s results, we recommend that China maintain its current statin initiation threshold. Although our findings suggest that relaxing the threshold would provide greater health economic benefits, clinical event results indicate that it would only marginally reduce the number of ASCVD events. ASCVD is a complex disease influenced by many factors and may not be best addressed solely through statins. Additionally, cost-effectiveness results show that a 7.5% 10-year ASCVD risk threshold would be more cost-effective with increased willingness to pay. However, China’s current economic level is not yet comparable to developed countries, and the average Chinese patient’s willingness to pay is low. Therefore, Chinese health policymakers should consider relaxing the treatment initiation threshold only after further economic improvement. Finally, hastily lowering the prophylaxis initiation threshold may cause a broken window effect, leading to increased use of other drugs such as anticoagulants and further burdening China’s healthcare system.

Markov model studies tend to have certain limitations. Firstly, Markov cohort models use average probabilities to model individual life courses, making it challenging to obtain more realistic simulation results[47]. In this study, we calculated the absolute ASCVD risk for each individual in the model based on demographic and clinical characteristics from the CHARLS study, allowing treatment decisions to be made based on individual risk. Secondly, a major characteristic of Markov cohort models is their memoryless nature, which also makes it difficult to model the impact of historical events on patient disease outcomes and costs[48]. Microsimulation models can address this by tracking each individual historically and adjusting individual transition probabilities and cost parameters across health states based on historical events.

This study inevitably has some unavoidable limitations. Firstly, the ASCVD risk function used in this study can only calculate the 10-year combined ASCVD risk, while the risks of developing AMI and stroke cannot be determined separately and must be simulated using a fixed probability level. Secondly, this study focused on the impact of statin-induced diabetes risk on ASCVD prevention and did not explore the health damage and economic loss due to complications in diabetic patients. This is a limitation of our study. Thirdly, previous studies have suggested that statin adherence significantly impacts ASCVD prevention. Due to the lack of data on long-term statin therapy in China, we used studies from other countries to model the impact of poor adherence, which may not accurately reflect the Chinese reality. Finally, although this study modeled the potential risk of statin-induced diabetes, it did not include all potential benefits and risks, such as reduced risk of advanced/aggressive prostate cancer, reduced mortality from in-hospital sepsis, and statin-induced rhabdomyolysis[49–52]. Previous studies have generally considered statin-induced diabetes to be the most significant negative scenario, so the potential factors not included in the analysis likely had minimal impact on this study[46].

### Conclusions

When willingness to pay is at 1-GDP, the current 10% 10-year ASCVD risk threshold used in China’s dyslipidemia management guidelines is cost-effective in preventing ASCVD events. Therefore, the current statin initiation threshold should be maintained. However, relaxing the 10-year ASCVD risk threshold would be a better strategy as willingness to pay increases. Changes in statin prices and the risk of statin-induced diabetes have a significant impact on cost-effectiveness outcomes.

## Supporting information

Supplemental Table 1, and will be used for the link to the file on the preprint site.

## Data Availability

Availability of data and materials
The datasets supporting the conclusions are available from the corresponding author (Yu X) on reasonable request.

## Abbreviations

AMI: Acute myocardial infarction
ASCVD: Atherosclerotic cardiovascular disease
CHD: Coronary heart disease
CHARLS: China Health and Retirement Longitudinal Study
China-PAR: Prediction for Atherosclerotic Cardiovascular Disease Risk in China
ICER: Incremental cost-benefit ratio
PSA: Probabilistic sensitivity analyses
QALY: Quality-adjusted life year
WTP: Willingness-to-pay

## Declarations

### Availability of data and materials

The datasets supporting the conclusions are available from the corresponding author (Yu X) on reasonable request.

### Consent for publication

Not applicable.

## Contributions

Tianyu Feng and Xihe Yu design research and draft paper. Data collection and analysis is done by Xiaolin Zhang and Jiaying Xu. Jiaying Xu and Shang Gao revised the article and put forward suggestions. All authors commented on previous versions of the manuscript. All authors read and approved the final manuscript.

## Acknowledgment

Not applicable.

## Ethics declarations

### Ethics approval and consent to participate

Not applicable.

## Consent for publication

Not applicable.

## References

1. Wang, L., et al., Prevalence and Treatment of Diabetes in China, 2013-2018. JAMA, 2021. 326(24): p. 2498–2506.

2. Zhou, M., et al., Cause-specific mortality for 240 causes in China during 1990-2013: a systematic subnational analysis for the Global Burden of Disease Study 2013. Lancet, 2016. 387(10015): p. 251–72.

3. Yang, G., et al., Rapid health transition in China, 1990-2010: findings from the Global Burden of Disease Study 2010. Lancet, 2013. 381(9882): p. 1987–2015.

4. Joint committee for guideline, r., 2016 Chinese guidelines for the management of dyslipidemia in adults. J Geriatr Cardiol, 2018. 15(1): p. 1–29.

5. Grundy, S.M., et al., 2018 AHA/ACC/AACVPR/AAPA/ABC/ACPM/ADA/AGS/APhA/ASPC/NLA/PCNA Guideline on the Management of Blood Cholesterol: A Report of the American College of Cardiology/American Heart Association Task Force on Clinical Practice Guidelines. Circulation, 2019. 139(25): p. e1082–e1143.

6. guideline CG181, N.I.C.E., Cardiovascular disease: risk assessment and reduction, including lipid modification. Methods. 2023, London.

7. Network, S.I.G., Risk estimation and the prevention of cardiovascular disease. 2007: Edinburgh: SIGN.

8. Cobiac, L.J., et al., Improving the cost-effectiveness of cardiovascular disease prevention in Australia: a modelling study. BMC Public Health, 2012. 12: p. 398.

9. Yang, X.L., et al., Risk stratification of atherosclerotic cardiovascular disease in Chinese adults. Chronic Dis Transl Med, 2016. 2(2): p. 102–109.

10. Casula, M., et al., Statin use and risk of new-onset diabetes: A meta-analysis of observational studies. Nutr Metab Cardiovasc Dis, 2017. 27(5): p. 396–406.

11. Mansi, I.A., et al., Association of Statin Therapy Initiation With Diabetes Progression: A Retrospective Matched-Cohort Study. JAMA Intern Med, 2021. 181(12): p. 1562–1574.

12. Galicia-Garcia, U., et al., Statin Treatment-Induced Development of Type 2 Diabetes: From Clinical Evidence to Mechanistic Insights. Int J Mol Sci, 2020. 21(13).

13. Li, Y., et al., Prevalence of diabetes recorded in mainland China using 2018 diagnostic criteria from the American Diabetes Association: national cross sectional study. BMJ, 2020. 369: p. m997.

14. Pandya, A., et al., Cost-effectiveness of 10-Year Risk Thresholds for Initiation of Statin Therapy for Primary Prevention of Cardiovascular Disease. JAMA, 2015. 314(2): p. 142–50.

15. MD, W. Population pyramids of the World from 1950 to 2100—China 2022. 2022.

16. Yang, X., et al., Predicting the 10-Year Risks of Atherosclerotic Cardiovascular Disease in Chinese Population: The China-PAR Project (Prediction for ASCVD Risk in China). Circulation, 2016. 134(19): p. 1430–1440.

17. Beck, J.R., et al., A convenient approximation of life expectancy (the “DEALE”). II. Use in medical decision-making. Am J Med, 1982. 73(6): p. 889–97.

18. Wu, Y., et al., Estimation of 10-year risk of fatal and nonfatal ischemic cardiovascular diseases in Chinese adults. Circulation, 2006. 114(21): p. 2217–25.

19. Greving, J.P., et al., Cost-effectiveness of aspirin treatment in the primary prevention of cardiovascular disease events in subgroups based on age, gender, and varying cardiovascular risk. Circulation, 2008. 117(22): p. 2875–83.

20. Yang, Z.J., et al., Prevalence of cardiovascular disease risk factor in the Chinese population: the 2007-2008 China National Diabetes and Metabolic Disorders Study. Eur Heart J, 2012. 33(2): p. 213–20.

21. Sattar, N., et al., Statins and risk of incident diabetes: a collaborative meta-analysis of randomised statin trials. Lancet, 2010. 375(9716): p. 735–42.

22. Brugts, J.J., et al., The benefits of statins in people without established cardiovascular disease but with cardiovascular risk factors: meta-analysis of randomised controlled trials. BMJ, 2009. 338: p. b2376.

23. Simons, L.A., et al., Discontinuation rates for use of statins are high. BMJ, 2000. 321(7268): p. 1084.

24. Simons, L.A., M. Ortiz, and G. Calcino, Persistence with antihypertensive medication: Australia-wide experience, 2004-2006. Med J Aust, 2008. 188(4): p. 224–7.

25. Poznyak, A., et al., The Diabetes Mellitus-Atherosclerosis Connection: The Role of Lipid and Glucose Metabolism and Chronic Inflammation. Int J Mol Sci, 2020. 21(5).

26. Laakso, M. and J. Kuusisto, Diabetes Secondary to Treatment with Statins. Curr Diab Rep, 2017. 17(2): p. 10.

27. Aiman, U., A. Najmi, and R.A. Khan, Statin induced diabetes and its clinical implications. J Pharmacol Pharmacother, 2014. 5(3): p. 181–5.

28. Organization, W.H., The global burden of disease: 2004 update. 2008: World Health Organization.

29. Tan, Z., et al., Health-related quality of life as measured with EQ-5D among populations with and without specific chronic conditions: a population-based survey in Shaanxi Province, China. PLoS One, 2013. 8(7): p. e65958.

30. Thakker, D., et al., Statin use and the risk of developing diabetes: a network meta-analysis. Pharmacoepidemiol Drug Saf, 2016. 25(10): p. 1131–1149.

31. Wang, S., et al., Direct medical costs of hospitalizations for cardiovascular diseases in Shanghai, China: trends and projections. Medicine (Baltimore), 2015. 94(20): p. e837.

32. Gu, D., et al., The Cost-Effectiveness of Low-Cost Essential Antihypertensive Medicines for Hypertension Control in China: A Modelling Study. PLoS Med, 2015. 12(8): p. e1001860.

33. Zhao, W., et al., Economic burden of obesity-related chronic diseases in Mainland China. Obes Rev, 2008. 9 Suppl 1: p. 62–7.

34. Sun, S., et al., Population health status in China: EQ-5D results, by age, sex and socio-economic status, from the National Health Services Survey 2008. Qual Life Res, 2011. 20(3): p. 309–20.

35. Wang, X., et al., Guideline for postmarketing Chinese medicine pharmacoeconomic evaluation. Chin J Integr Med, 2015. 21(6): p. 473–80.

36. Robinson, L.A., et al., Understanding and improving the one and three times GDP per capita cost-effectiveness thresholds. Health Policy Plan, 2017. 32(1): p. 141–145.

37. Bleibler, F., et al., Expected lifetime numbers and costs of fractures in postmenopausal women with and without osteoporosis in Germany: a discrete event simulation model. BMC Health Serv Res, 2014. 14: p. 284.

38. Eddy, D.M., et al., Model transparency and validation: a report of the ISPOR-SMDM Modeling Good Research Practices Task Force-7. Med Decis Making, 2012. 32(5): p. 733–43.

39. Vemer, P., et al., AdViSHE: A Validation-Assessment Tool of Health-Economic Models for Decision Makers and Model Users. Pharmacoeconomics, 2016. 34(4): p. 349–61.

40. Wang, W., et al., Prevalence, Incidence, and Mortality of Stroke in China: Results from a Nationwide Population-Based Survey of 480 687 Adults. Circulation, 2017. 135(8): p. 759–771.

41. Boone, J., Basic versus supplementary health insurance: Access to care and the role of cost effectiveness. J Health Econ, 2018. 60: p. 53–74.

42. Collins, R., et al., Interpretation of the evidence for the efficacy and safety of statin therapy. Lancet, 2016. 388(10059): p. 2532–2561.

43. Simic, I. and Z. Reiner, Adverse effects of statins - myths and reality. Curr Pharm Des, 2015. 21(9): p. 1220–6.

44. Rosenbaum, L., Beyond belief--how people feel about taking medications for heart disease. N Engl J Med, 2015. 372(2): p. 183–7.

45. Xiong, X., et al., Impact of universal medical insurance system on the accessibility of medical service supply and affordability of patients in China. PLoS One, 2018. 13(3): p. e0193273.

46. Jiang, Y. and W. Ni, Economic Evaluation of the 2016 Chinese Guideline and Alternative Risk Thresholds of Initiating Statin Therapy for the Management of Atherosclerotic Cardiovascular Disease. Pharmacoeconomics, 2019. 37(7): p. 943–952.

47. Caro, J.J., J. Moller, and D. Getsios, Discrete event simulation: the preferred technique for health economic evaluations? Value Health, 2010. 13(8): p. 1056–60.

48. Hiligsmann, M., et al., Development and validation of a Markov microsimulation model for the economic evaluation of treatments in osteoporosis. Value Health, 2009. 12(5): p. 687–96.

49. Boudreau, D.M., O. Yu, and J. Johnson, Statin use and cancer risk: a comprehensive review. Expert Opin Drug Saf, 2010. 9(4): p. 603–21.

50. Ouellette, D.R., et al., Sepsis outcomes in patients receiving statins prior to hospitalization for sepsis: comparison of in-hospital mortality rates between patients who received atorvastatin and those who received simvastatin. Ann Intensive Care, 2015. 5: p. 9.

51. Mendes, P., P.G. Robles, and S. Mathur, Statin-induced rhabdomyolysis: a comprehensive review of case reports. Physiother Can, 2014. 66(2): p. 124–32.

52. Ganda, O.P., Statin-induced diabetes: incidence, mechanisms, and implications. F1000Res, 2016. 5

