## Supplemental Table 1, and will be used for the link to the file on the preprint site. for "Health Economics Assessment of Statin Therapy Initiation Thresholds for Atherosclerosis Prevention in China: a Cost-Effectiveness Analysis"

Table S1. Age-specific cohort risk factor distributions. SBP and WC data were from the 2015 wave of CHARLS data. HDL-C, LDL-C.

|  | SBP  (mmHg) | | WC  (cm) | | HDL-C  (mg/dL) | | TG  (mg/dL) | | LDL  (mg/dL) | |
| --- | --- | --- | --- | --- | --- | --- | --- | --- | --- | --- |
| Age group | Mean | SD | Mean | SD | Mean | SD | Mean | SD | Mean | SD |
| Male |  |  |  |  |  |  |  |  |  |  |
| 45-49 | 123.7 | 12.1 | 86.9 | 12.1 | 47.1 | 36.2 | 145.1 | 136.3 | 114.0 | 36.2 |
| 50-54 | 123.7 | 10.9 | 87.7 | 10.9 | 48.2 | 34.4 | 148.6 | 136.7 | 111.9 | 34.4 |
| 55-59 | 126.1 | 11.8 | 87.2 | 11.8 | 49.7 | 34.3 | 127.3 | 110.5 | 114.4 | 34.3 |
| 60-64 | 128.0 | 12.0 | 86.0 | 12.0 | 49.4 | 35.8 | 130.8 | 105.9 | 112.9 | 35.8 |
| 65-69 | 127.9 | 13.7 | 86.0 | 13.7 | 50.1 | 33.0 | 122.7 | 85.9 | 110.7 | 33.0 |
| 70-74 | 129.2 | 12.8 | 84.2 | 12.8 | 50.4 | 30.2 | 115.0 | 94.4 | 110.3 | 30.2 |
| 75-79 | 130.3 | 14.5 | 84.2 | 14.5 | 50.6 | 30.9 | 103.4 | 58.4 | 110.0 | 30.9 |
| 80-84 | 138.9 | 14.5 | 83.5 | 14.5 | 52.4 | 30.5 | 92.3 | 58.8 | 98.6 | 30.5 |
| 85-89 | 134.2 | 13.5 | 82.7 | 13.5 | 44.8 | 23.7 | 136.4 | 74.1 | 95.5 | 23.7 |
| Female |  |  |  |  |  |  |  |  |  |  |
| 45-49 | 117.4 | 12.5 | 84.8 | 12.5 | 50.6 | 31.1 | 127.1 | 90.1 | 110.9 | 31.1 |
| 50-54 | 120.5 | 10.6 | 86.1 | 10.6 | 49.6 | 35.1 | 139.1 | 108.4 | 119.3 | 35.1 |
| 55-59 | 121.8 | 11.5 | 86.0 | 11.5 | 50.7 | 34.2 | 140.4 | 97.1 | 121.2 | 34.2 |
| 60-64 | 122.8 | 13.0 | 86.4 | 13.0 | 49.2 | 36.9 | 145.7 | 103.3 | 120.8 | 36.9 |
| 65-69 | 128.4 | 13.0 | 86.2 | 13.0 | 48.5 | 36.3 | 163.0 | 141.0 | 118.3 | 36.3 |
| 70-74 | 131.2 | 13.4 | 86.6 | 13.4 | 49.9 | 36.8 | 141.5 | 107.5 | 124.1 | 36.8 |
| 75-79 | 135.3 | 13.8 | 85.9 | 13.8 | 50.5 | 38.3 | 132.5 | 75.2 | 119.9 | 38.3 |
| 80-84 | 136.6 | 14.5 | 86.3 | 14.5 | 55.2 | 36.0 | 144.7 | 84.2 | 119.3 | 36.0 |
| 85-89 | 134.7 | 10.4 | 84.4 | 10.4 | 50.0 | 34.9 | 123.2 | 74.7 | 116.6 | 34.9 |

Table S2. Age-specific background mortality and utility inputs

| Input type | value | | References |
| --- | --- | --- | --- |
| Annual background mortality | Men | Women | [1] |
| Age 45–49 | 0.059% | 0.043% |  |
| Age 50–54 | 0.099% | 0.070% |  |
| Age 55–59 | 0.176% | 0.119% |  |
| Age 60–64 | 0.307% | 0.211% |  |
| Age 65–69 | 0.527% | 0.373% |  |
| Age 70–74 | 0.894% | 0.653% |  |
| Age 75–79 | 1.484% | 1.128% |  |
| Age 80–84 | 2.600% | 2.063% |  |
| Age 85–89 | 4.223% | 3.512% |  |
| Age-specific utility | Men | Women | [1] |
| Age 45–49 | 0.814 | 0.792 |  |
| Age 50–54 | 0.793 | 0.772 |  |
| Age 55–59 | 0.774 | 0.752 |  |
| Age 60–64 | 0.751 | 0.728 |  |
| Age 65–69 | 0.725 | 0.702 |  |
| Age 70–74 | 0.701 | 0.685 |  |
| Age 75–79 | 0.684 | 0.669 |  |
| Age 80–84 | 0.662 | 0.655 |  |
| Age 85–89 | 0.661 | 0.643 |  |

Table S3. Age distributions by sex.

| Age group | Male | Female |
| --- | --- | --- |
| 45-49 | 17.27% | 21.75% |
| 50-54 | 17.59% | 16.57% |
| 55-59 | 15.00% | 13.69% |
| 60-64 | 17.05% | 16.25% |
| 65-69 | 12.98% | 10.89% |
| 70-74 | 8.26% | 7.96% |
| 75-79 | 5.86% | 4.97% |
| 80-84 | 3.25% | 3.18% |
| 85-89 | 2.74% | 4.73% |

Table S4. Distributions of smoking, disease history, treatment history and regional.

|  | Male | Female |
| --- | --- | --- |
| Smoking | 53.77% | 5.11% |
| Diabetes | 13.4% | 13.0% |
| Northern | 40.82% | 40.82% |
| Urban residence | 49.29% | 49.29% |
| Family history of ASCVD | 13.4% | 13.0% |

Table S5. Relationship between risk factors and age based on linear regressions using CHARLS data.

| Risk factor | Male | Female |
| --- | --- | --- |
| SBP^a^ | $SBP=106.72+0.3683\times age$ | $SBP=85.845+0.6916\times age$ |
| WC | $WC=85.631+0.0131\times age$ | $WC=84.812+0.0244\times age$ |
| HDL-C^a^ | $HDL-C=47.224+0.0461\times age$ | $HDL-C=49.9+0.0397\times age$ |
| LDL-C | $LDL-C=2.5370+0.0713\times age$ | LDL-C= 2.6411 + 0.0682×age |
| TG^a^ | $TG=190.88-0.2157\times age$ | $TG=155.34+0.5699\times age$ |

Table S6. Calculation equations of Risk of CVD base on China-PAR Risk Function.

MALE

31.97×ln(age)+0.62×ln(TC)-0.69×ln(HDL-C)-0.71×ln(waist) (+[27.39-6.02×ln(age)] ×ln(SBP) if hypertension treated) (+[26.15-5.73×ln(age)]×ln(SBP) if hypertension untreated) (+3.96-0.94×ln(age) if current smoker) (+6.22-1.53×ln(age) if having family history of ASCVD) (+0.36 if diabetes) (+0.48 if in Northern China) (-0.16 if living in urban)

FEMALE

24.87×ln(age)+0.06×ln(TC)-0.22×ln(HDL-C)+1.48×ln(waist) (+[20.71-4.53×ln(age)] ×ln(SBP) if hypertension treated) (+[19.98 -4.36×ln(age)]×ln(SBP) if hypertension untreated) (+0.49 if current smoker) (+0.57 if diabetes) (+0.54 if in Northern China)

China-PAR risk estimation was calculated as: Predicted ASCVD risk = 1 - S0(t)^exp(Individual score – Mean score).

Fig S1. Illustration of the strategy of triggering statin therapy based on the 2023 Chinese guidelines for the management of dyslipidemia in adults.

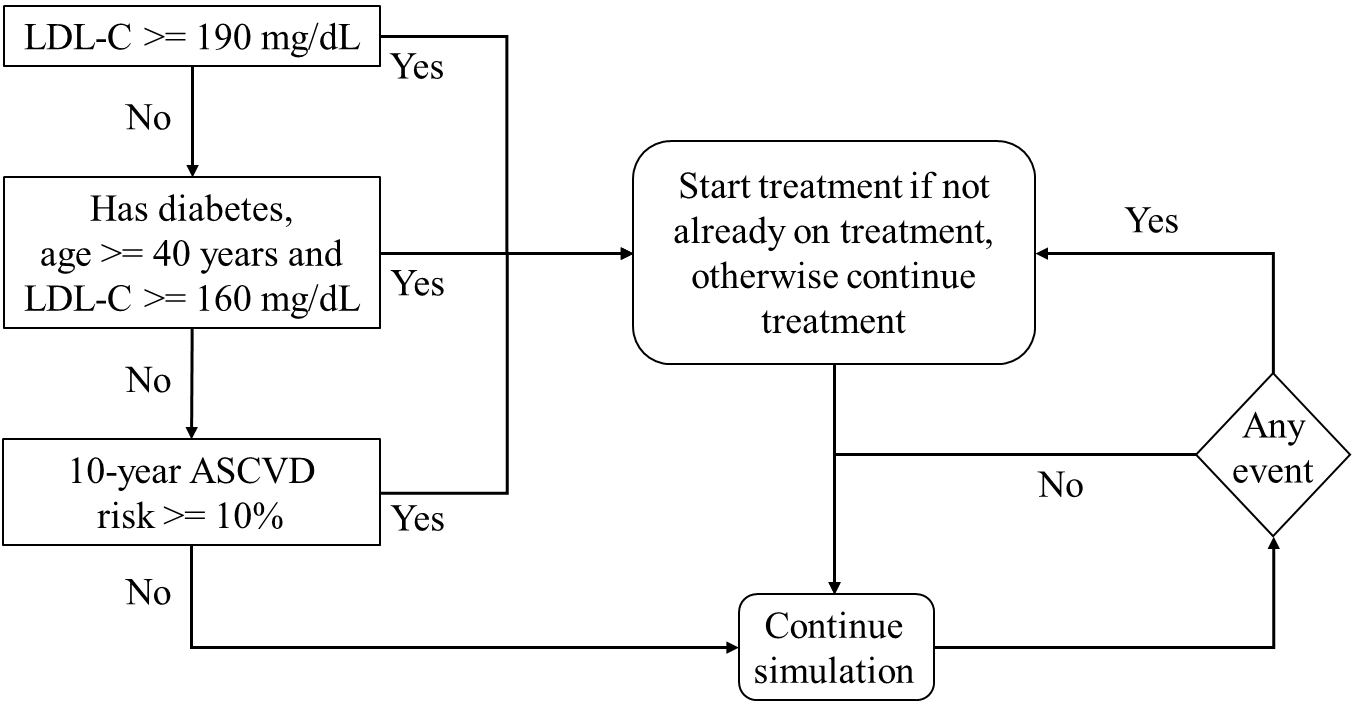

Table S7. Incidence rates of stroke per 100,000 person-years of Chinese adults from the current simulation study and an independent prospective study in China [2] for the examination of the external validity of the model.

|  | Age group | Simulated | Reference study |
| --- | --- | --- | --- |
| Male | 50-59 | 404.8 | 528.5 |
|  | 60-69 | 802.6 | 908.6 |
|  | 70-79 | 1187.6 | 1486.9 |
|  | 80+ | 1994.6 | 2216.6 |
| Female | 50-59 | 161.8 | 433.1 |
|  | 60-69 | 373.5 | 821.4 |
|  | 70-79 | 627.9 | 1349.9 |
|  | 80+ | 1135.6 | 2095.8 |

Fig S2. Expected vs. simulated percentages of patients that had CHD and stroke in a year for selected age-sex groups (age in multiple of ten) for the examination of the internal validity of the model. The expected values were calculated using the risk equation and the means of the risk factor distributions in the input data. The R^2^ of the regression of the expected values on the simulated values was 0.9278.

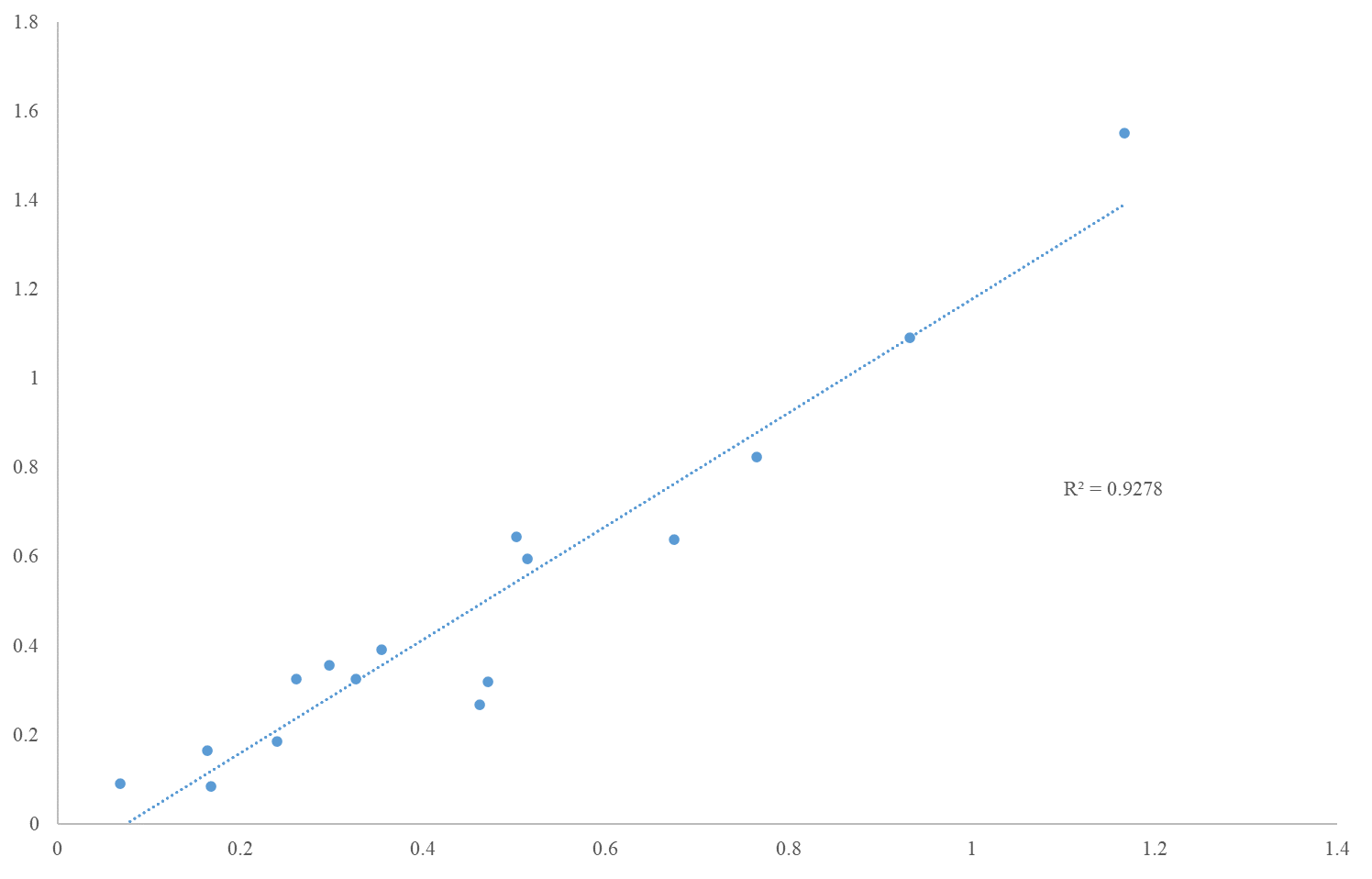

**Reference**

1.	Sun, S., et al., *Population health status in China: EQ-5D results, by age, sex and socio-economic status, from the National Health Services Survey 2008.* Qual Life Res, 2011. **20**(3): p. 309-20.

2.	Wang, W., et al., *Prevalence, Incidence, and Mortality of Stroke in China: Results from a Nationwide Population-Based Survey of 480 687 Adults.* Circulation, 2017. **135**(8): p. 759-771.
